# Testing for Causal Association between Serum Urate, Gout, and Prostatic Cancer in European Males

**DOI:** 10.1101/2025.05.09.25327351

**Authors:** Sumanth R. Chandrupatla, Nicholas Sumpter, Riku Takei, Tony R. Merriman, Jasvinder A. Singh

**Affiliations:** Heersink School of Medicine, University of Alabama at Birmingham, AL, United States; Division of Clinical Immunology and Rheumatology, University of Alabama at Birmingham, AL, United States; Department of Internal Medicine, Radboud University Medical Center, Nijmegen, Netherlands; Medicine Service, Michael E. DeBakey VA Medical Center, 2002 Holcombe Blvd, Houston, TX 77030, USA; Department of Medicine, Baylor College of Medicine, 7200 Cambridge Street, Houston, TX 77030, USA

## Abstract

**Objective:** To conduct a two-sample MR study including only men to test for a causal relationship between serum urate (SU) and gout and prostate cancer.

**Methods:** We used GWAS for SU, gout, and prostate cancer to generate exposure instrumental variables (IV) associated with gout and urate. We used 20 single nucleotide polymorphisms (SNPs) associated with gout but not urate for an IV representing the non-hyperuricemia (inflammatory) compartment of gout and we used four SNPs from loci containing urate transporter genes for an IV representing urate levels. MR methods included inverse-variance-weighted (IVW), MR-Egger regression, and weighted median to test for causal relationships and horizontal pleiotropy.

**Results:** The non-hyperuricemia compartment of gout IV showed a causal effect of gout on prostate cancer (Weighted median: *P* = 0.01). In contrast, the SU IV showed no evidence for a causal effect of SU on prostate cancer (IVW: *P* = 0.83; Weighted Median: *P* = 0.97). We found no evidence of horizontal pleiotropy from MR-Egger for either the urate or gout IV (non-hyperuricemia compartment of gout: *P* = 0.33; urate: *P* = 0.80). Loci contributing most strongly to the non-hyperuricemia causal effect included three genes: *IL1R1, IL1RN*, and *SLC30A5*. There was no evidence for a causal relationship of prostate cancer on gout or SU.

**Conclusion:** MR analysis in a European male population found evidence for a causal effect between the non-hyperuricemia compartment of gout and prostate cancer. Implication of the *IL1R1* and *IL1RN* genes directly implicates the gouty inflammation pathway in prostate cancer.

## Introduction

Gout is an inflammatory disorder characterized by elevation of SU that manifests as local and systemic inflammation secondary to monosodium urate (MSU) crystal deposition in joints and other tissues. If left untreated, gout frequently manifests as recurrent episodes of acute inflammation that are very painful and severely disabling, potentially increasing the risk of comorbidities including heart failure, myocardial infarction, chronic kidney disease stage ≥2 and diabetes (1-3). Gout affects approximately 5.1% of adults in the US, with ∼3-fold increased prevalence in men, and men have elevated SU levels compared to women (4-7). There is evidence that the prostate plays a role in urate homeostasis in men, with urate being present in human semen, and urate crystals being found in 47.5% of non-malignant prostate sections (8-10). Additionally, gout is associated with an increased risk of benign prostatic hyperplasia (BPH) and treatment with the urate-lowering drug allopurinol decreases the risk of BPH, indicating that the presence of higher urate levels and gout may both contribute to prostatic enlargement (11, 12). BPH shares a number of features with prostate cancer, including hormone-dependent growth and metabolic disruption playing key roles in disease development, and a causal relationship between BPH and prostate cancer has recently been established (13, 14).

Mendelian randomization (MR) is a genetic epidemiological approach to establish causality between two phenotypes by testing genetic variants (instrumental variable) associated with increased risk of the exposure with the outcome (17). One advantage of using MR is that genetic variants are fixed at conception, making the method robust to the influence of confounding factors (15, 16). Previous studies using MR examined whether a causal relationship exists between urate and prostate cancer (17, 18), though none have tested the association of gout with prostate cancer. Studies that have examined genetic links between SU and prostate cancer have had contradictory findings. In an East Asian cohort, increased urate levels were causally associated with prostate cancer, but no causal relationship was found in a European cohort (17, 18). However, these studies included females in the GWAS for development of the SU instrumental variable. Since females have lower SU levels (5), and prostate cancer only affects males, the results found from these studies may be biased.

Due to the genetic and epidemiological evidence of a potential relationship between gout, urate, prostatic inflammation, and prostate cancer, limitations with previous MR studies, and the fact that gout has not yet been examined as an exposure in prostate cancer MR, we aimed to identify whether there was a potential causal relationship between urate, gout-related inflammation, and prostate cancer. Unlike all previous MR studies in gout, we isolated the non-hyperuricemic (inflammatory) compartment of gout and tested this signal along with urate transport genes against prostate-cancer risk.

## Methods

### Exposure instrumental variable and outcome GWAS summary statistics for two-sample MR study

Our study was designed to identify any potential causal effects of SU and of the non-hyperuricemia compartment of gout on prostate cancer risk. Since prostate cancer is a male-only condition, we identified male-only genome-wide association studies (GWAS) of SU and gout, and used effect sizes from these studies in the MR analysis. We used previously published European ancestry male-only gout and SU GWAS (19). The gout GWAS contained 77,628 cases and 933,894 controls, and the SU GWAS contained a total of 145,625 men aged 40-69 with complete SU measurements from the UK Biobank (project number 12611). The prostate cancer GWAS was sourced from Wang, et al. and contained 122,188 cases and 604,640 male controls of European ancestry (20).

### Statistical Analysis

We selected gout instrumental variables (IVs) from the gout GWAS to represent the non-hyperuricemia compartment of gout. This was determined previously by Major, et al. by conducting colocalization analysis between the lead variants for signals in gout and SU (19), where lead SNPs from signals with posterior probabilities for H1 or H3 ≥ 0.8 represented an association with gout but not urate, and therefore the non-hyperuricemia compartment of gout (such as promotion of gout inflammation and deposition of MSU crystals). The lead SNPs from these genetic signals were used for MR analysis as IVs (19). SU IVs were selected as the lead SNPs from the SU GWAS at loci including genes involved in urate transport: *SLC2A9, SLC2A11, PDZK1, SLC17A1*. This was done to minimize pleiotropy owing to the established function of the four proteins in directly influencing urate levels and without robust evidence for a role in other physiological processes. F-statistics were calculated for each SNP from both the gout and SU GWAS, as described previously (21). A full list of all SNPs used for the non-hyperuricemia compartment of gout, all proxy variants selected, and a list of all SNPs used to represent urate transporter genes are available in **Supplementary Table 1**. Proxy variants were selected when a SNP in the IV was not present in the prostate cancer GWAS. Proxies were selected by identifying variants in high LD (r^2^ > 0.95) and selecting the top variant available in both the SU/gout and prostate cancer GWAS,

We also conducted the reverse MR, i.e. testing prostate cancer for causality of gout and SU. Due to the large number of significant risk variants in the source prostate cancer GWAS (451 risk variants were identified by Wang, et al.) (20), we selected only the most significant SNPs (*P* < 1e-80) to reduce the risk of pleiotropy. The IV was identified by first identifying loci with SNPs associated with prostate cancer at a level of *P* < 1e-50. After these loci were identified, we selected only the top SNP from each locus, keeping only SNPs with *P* < 1e-80. This resulted in a set of 18 SNPs (**Supplementary Table 2**).

For these IVs to be valid, they each should fulfill the assumptions necessary for MR: [1] all selected IVs are significantly associated with the exposure (F-Statistic > 10); [2] IVs are independent of confounders that could affect both the exposure and outcome; [3] IVs influence the outcome only through the exposure (15). We did not evaluate assumption 2 because we did not identify any obvious conditions that may be potential confounders or mediators between gout/SU and prostate cancer. Assumption 3 was tested using MR-Egger, which accounts for potential horizontal pleiotropic effects between the exposure and outcomes (15).

Three main methods of MR were used: inverse-variance-weighted MR (IVW), MR-Egger regression, and weighted median. Additional sensitivity analyses included penalized IVW and the exclusion of outlier SNPs. All MR analyses used R 4.3.1, and the MendelianRandomization package (22). For all MR analyses, we used the effect size for the outcome as the response variable and the effect size for the exposure as the predictor variable. This resulted in two main relationships being tested: [1] the non-hyperuricemia compartment of gout to be causal of prostate cancer; [2] SU to be causal of prostate cancer. The reverse relationships were also tested: [1] prostate cancer to be causal of gout; and [2] prostate cancer to be causal of SU.

After MR analysis, we examined whether the SNPs driving the MR results acted as potential expression quantitative trait loci (eQTL) in the following tissues from the Genotype-Tissue Expression (GTEx) data (22): whole blood, prostate, cultured fibroblast cells, Epstein-Barr-virus (EBV)-transformed lymphocytes, and testis. The data for these analyses were obtained from dbGaP accession number phs000424.v10.p2, and European GTEx v8 eQTL data were used. Colocalization results were sourced from Major et al (19).

## Results

### Gout to prostate cancer Mendelian randomization

Using the weighted median method we observed evidence that gout may be causal of prostate cancer (OR: 1.18; 95% CI: 1.03-1.35; P=0.01; Table 1; Figure 1) although this was not supported using the IVW method (IVW: OR: 1.10; 95% CI: 0.92-1.31; P=0.29; Table 1; Figure 1). MR-Egger showed no evidence of horizontal pleiotropy (Intercept: -0.010; SE: 0.010; P=0.33; Table 1). Sensitivity analysis using the penalized IVW method to handle potential outliers supported our hypothesis that gout is causal of prostate cancer (OR: 1.16; 95% CI: 1.03-1.31; P=0.01; **Supplementary Table 3**). The result of all gout non-hyperuricemia compartment SNPs is available in **Supplementary Figure 1**. The gout risk alleles of three SNPs, rs2560449, rs17767183, and rs9973741, drove this causal relationship, while the gout risk allele of one SNP, rs2395180, was significantly protective of prostate cancer.

**Table 1:**
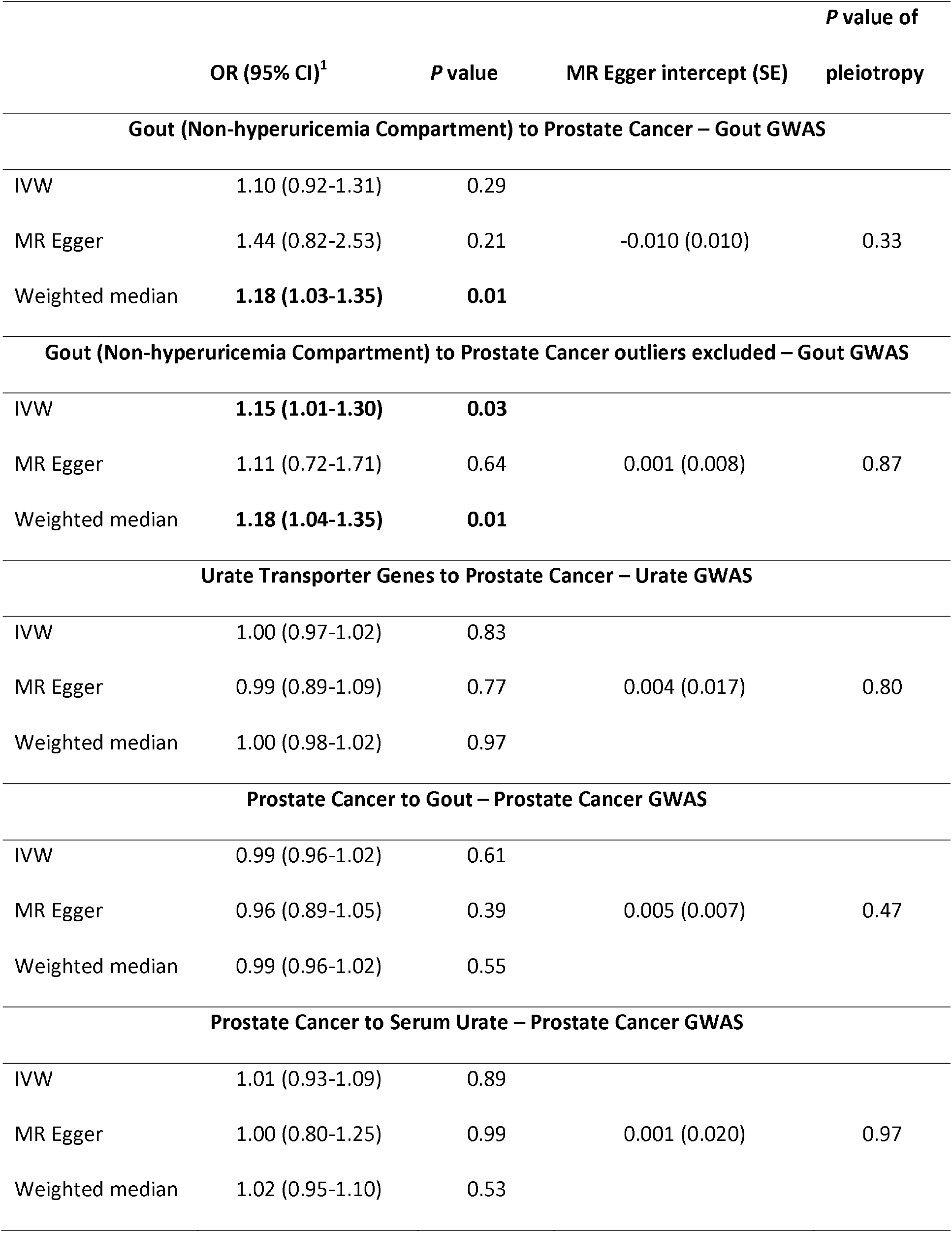
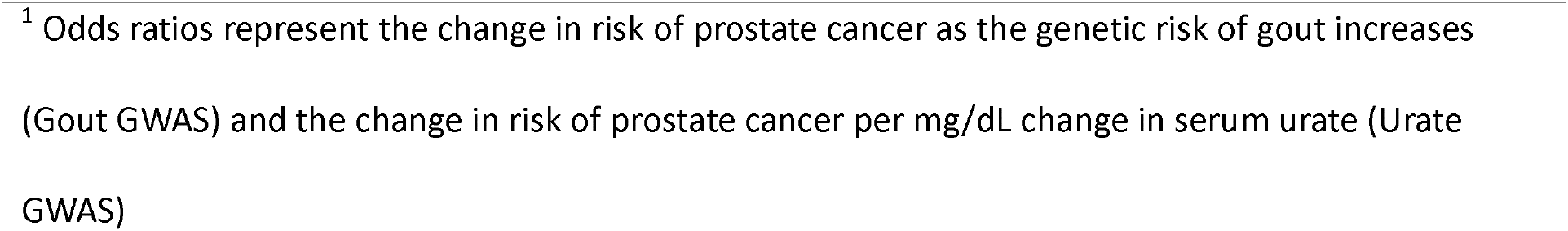
Mendelian Randomization Results.

|  | OR (95% CI)<sup>1</sup> | P value | MR Egger intercept (SE) | P value of pleiotropy |
| --- | --- | --- | --- | --- |
| **Gout (Non-hyperuricemia Compartment) to Prostate Cancer – Gout GWAS** |  |  |  |  |
| IVW | 1.10 (0.92-1.31) | 0.29 |  |  |
| MR Egger | 1.44 (0.82-2.53) | 0.21 | -0.010 (0.010) | 0.33 |
| Weighted median | **1.18 (1.03-1.35)** | **0.01** |  |  |
| **Gout (Non-hyperuricemia Compartment) to Prostate Cancer outliers excluded – Gout GWAS** |  |  |  |  |
| IVW | **1.15 (1.01-1.30)** | **0.03** |  |  |
| MR Egger | 1.11 (0.72-1.71) | 0.64 | 0.001 (0.008) | 0.87 |
| Weighted median | **1.18 (1.04-1.35)** | **0.01** |  |  |
| **Urate Transporter Genes to Prostate Cancer – Urate GWAS** |  |  |  |  |
| IVW | 1.00 (0.97-1.02) | 0.83 |  |  |
| MR Egger | 0.99 (0.89-1.09) | 0.77 | 0.004 (0.017) | 0.80 |
| Weighted median | 1.00 (0.98-1.02) | 0.97 |  |  |
| **Prostate Cancer to Gout – Prostate Cancer GWAS** |  |  |  |  |
| IVW | 0.99 (0.96-1.02) | 0.61 |  |  |
| MR Egger | 0.96 (0.89-1.05) | 0.39 | 0.005 (0.007) | 0.47 |
| Weighted median | 0.99 (0.96-1.02) | 0.55 |  |  |
| **Prostate Cancer to Serum Urate – Prostate Cancer GWAS** |  |  |  |  |
| IVW | 1.01 (0.93-1.09) | 0.89 |  |  |
| MR Egger | 1.00 (0.80-1.25) | 0.99 | 0.001 (0.020) | 0.97 |
| Weighted median | 1.02 (0.95-1.10) | 0.53 |  |  |

<sup>1</sup> Odds ratios represent the change in risk of prostate cancer as the genetic risk of gout increases (Gout GWAS) and the change in risk of prostate cancer per mg/dL change in serum urate (Urate GWAS)

**Figure 1:**
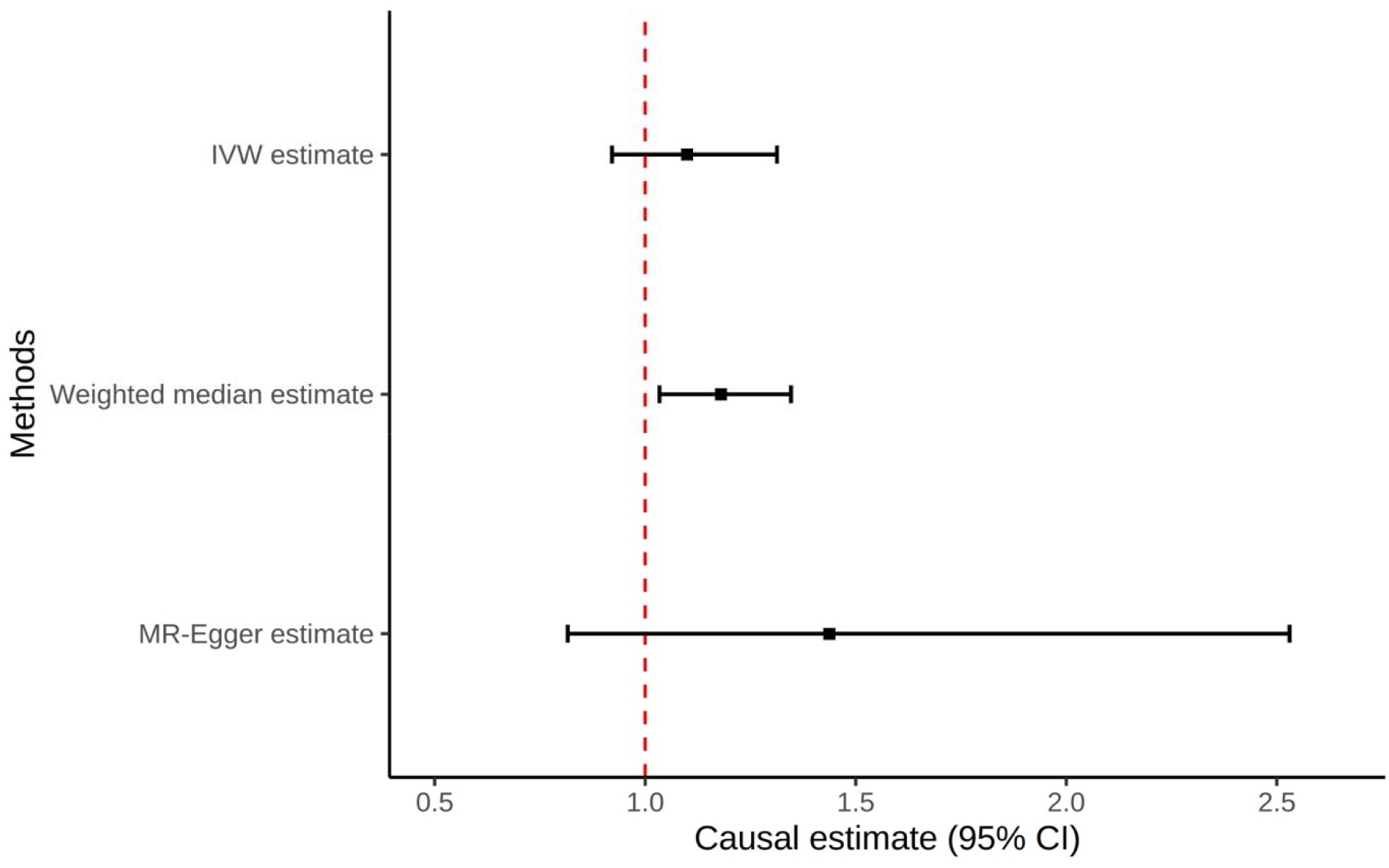
Forest plot of all Mendelian Randomization tests for the non-hyperuricemia compartment of gout to prostate cancer

LocusZoom plots were generated for each SNP that significantly contributed to the positive association between gout and prostate cancer, and each SNP was assigned to the closest gene: *SLC30A5* (rs2560449; **Supplementary Figure 2**), *IL1R1* (rs17767183; **Supplementary Figure 3**), *IL1RN* (rs9973741; **Supplementary Figure 4**). Even though rs9973741 is in very close proximity to both *IL1RN* and *IL1F10*, we only assigned it to *IL1RN* since *IL1F10* is only expressed in skin tissue (GTEx). We noted that rs2395180 was located in the *HLA* locus, in close proximity to a number of genes (**Supplementary Figure 5**). Using the Major et al. data (19) we identified one potentially colocalizing eQTL from the SNPs and tissues of interest: rs9973741 with *IL1RN* expression in the testis (**Supplementary Table 4; Supplementary Figure 6**). The *IL1RN* eQTL signal overlaps only with gout, and not with prostate cancer (**Supplementary Figure 6**).

We conducted a secondary analysis excluding rs2395180 as a clear outlier with non-overlapping 95% confidence intervals (CI) with other variants in the IV and being in a locus with erratic association with gout, perhaps driven by (uncorrected) population structure (23). In this analysis, IVW, weighted median, and penalized IVW methods all showed causal effects of gout on prostate cancer (**Supplementary Table 3**).

### Serum urate to prostate cancer Mendelian randomization

Using the IVW method, there was no evidence that genetically predicted SU was causal of prostate cancer (OR: 1.00; 95% CI: 0.97-1.02; *P* = 0.83; **Table 1; Figure 3**). This lack of causal relationship was also seen in the weighted median analysis (OR: 1.00; 95% CI: 0.98-1.02; P=0.97; **Table 1; Figure 3**). Additionally, MR-Egger showed no evidence for horizontal pleiotropy (Intercept: 0.004; SE: 0.017; P=0.80; **Table 1**). No individual SNPs were significantly associated.

**Figure 2:**
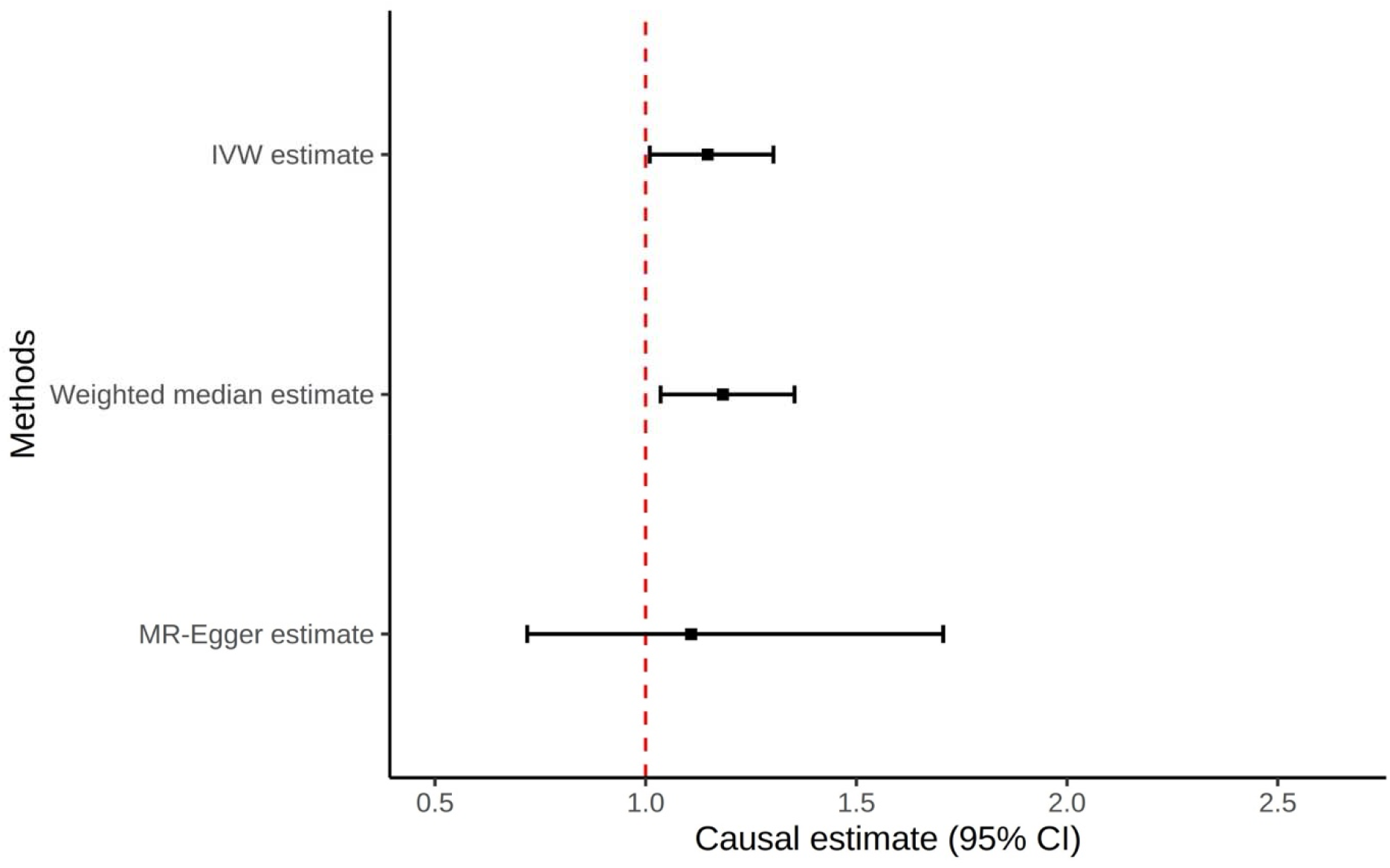
Forest plot of all Mendelian Randomization tests for the non-hyperuricemia compartment of gout to prostate cancer, excluding outlier (rs2395180)

**Figure 3:**
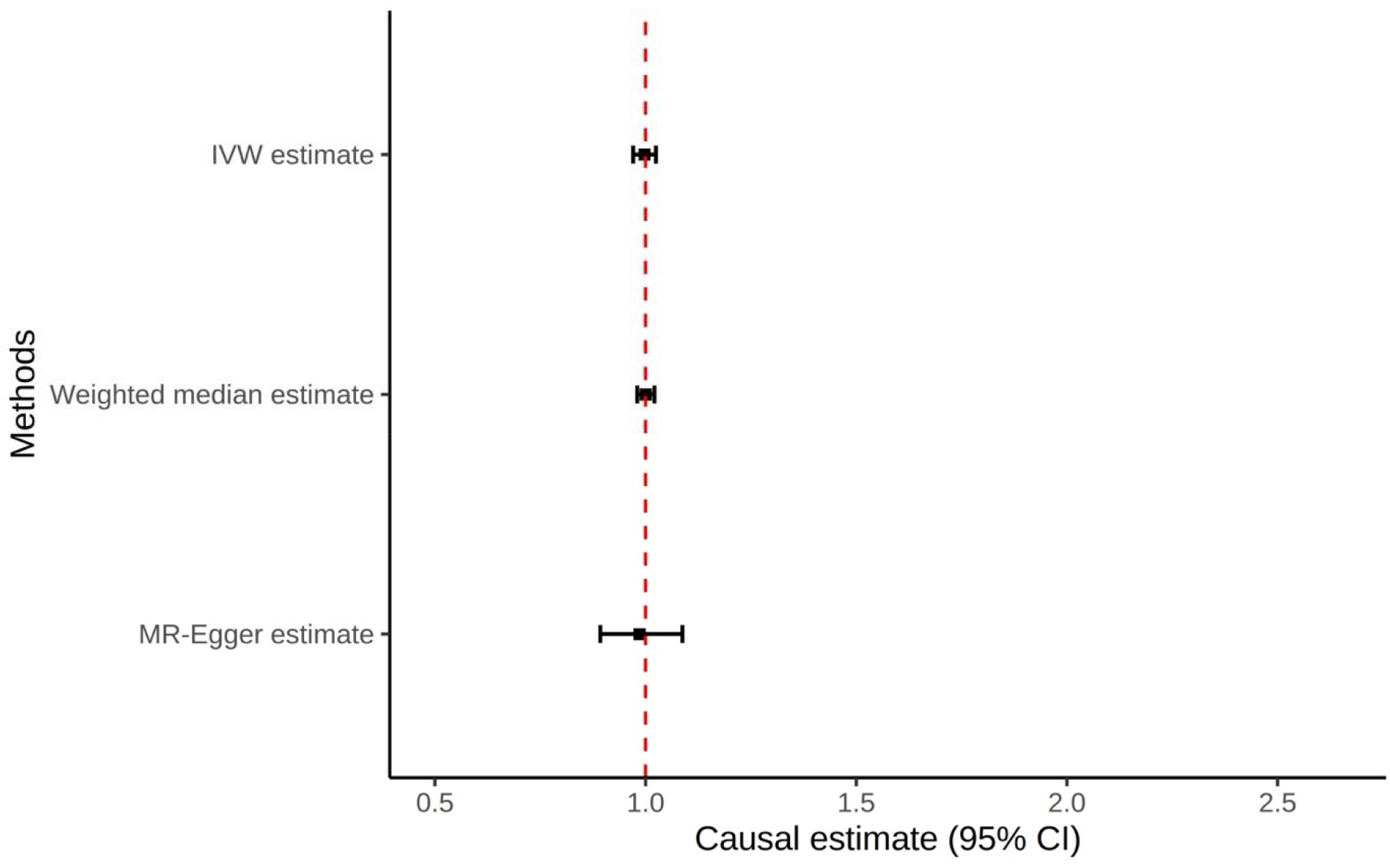
Forest plot of all Mendelian Randomization tests for the urate transporter SNPs to prostate cancer

### Prostate cancer to serum urate and gout Mendelian randomization

There was no evidence that genetically predicted prostate cancer was causal of gout using either the IVW (OR: 0.99; 95% CI: 0.96-1.02; *P* = 0.61; **Table 1**) or weighted median MR methods (OR: 0.99; 95% CI: 0.96-1.02; *P* = 0.53; **Table 1**). There was also no evidence that genetically predicted prostate cancer was causal of SU using either the IVW method (OR: 1.01; 95% CI: 0.93-1.09; *P* = 0.89; **Table 1**) or the weighted median method (OR: 1.02; 95% CI: 0.95-1.10; *P* = 0.53; **Table 1**). Additionally, MR-Egger showed no evidence for horizontal pleiotropy in either gout (Intercept: 0.005; SE: 0.007; P=0.47; **Table 1**) or SU (Intercept: 0.001; SE: 0.020; P=0.97; **Table 1**).

## Discussion

Our study tested whether the non-hyperuricemia compartment of gout and SU are causal of prostate cancer in men of European ancestry. We assessed this using a two-sample MR analysis in men and found evidence supporting a causal relationship between gout inflammation (two of the three positively association variants were in loci encoding genes central to IL-1β signaling) and prostate cancer. There was no evidence for a causal relationship for SU on prostate cancer, or for prostate cancer on gout or SU.

Our findings align with previous MR findings that SU is not causal of prostate cancer among European males (18), although there have been mixed findings from MR studies in East Asians and in observational studies, with observational studies showing differences in SU levels in those with prostate cancer vs those without (17, 24-26). However, none of these studies assessed the reverse relationships or gout-causing inflammatory factors, and observational studies cannot infer causality.

To our knowledge this is the first evidence that the non-hyperuricemia compartment of gout may be causal of prostate cancer. There have been observational studies of gout and prostate cancer, but the results have been conflicting. In one cohort study of Taiwanese patients with 8,408 male gout patients and 25,010 age-matched and time-of-diagnosis-matched controls, gout patients were 3 times more likely to develop prostate cancer when compared to the non-gout population (27). In another Taiwanese cohort study including 355,278 male patients, 25,943 of whom had gout, gout patients were 1.7 times more likely to develop prostate cancer than non-gout patients (28). However, in a Korean health insurance data study of 179,930 age-matched gout patients and non-gout controls, men with prostate cancer had a lower incidence rate of gout when compared to age-matched non-gout controls (29). In another study of hospital discharges in Sweden, there was no association between gout and the incidence of prostate cancer (30).

Our MR study identified three genes driving the relationship between the non-hyperuricemia compartment of gout and prostate cancer: *IL1R1* (rs17767183), *IL1RN* (rs9973741), and *SLC30A5* (rs2560449). *IL1R1* (31), *Il1RN* (32), and *SLC30A5* (33) have all been previously implicated in prostate cancer. The IL1 pathway has been shown to play a role in both the development and progression of prostate cancer, though the exact mechanism is still unknown (31, 32). Additionally, depleted zinc concentration is associated with prostate cancer progression, and zinc transporter *SLC30A5* has been shown to be downregulated in cancerous tissue (33). Though we do not fully understand the mechanisms via which these SNPs may be causal of prostate cancer, it is notable that two of the top SNPs are both involved in the same inflammatory pathway (the IL1 pathway) with the protein encoded by *IL1RN* (IL1-RA) being an antagonist for IL1-R1 which is also a receptor for the key gout cytokine IL-1β. This biological linkage supports the hypothesis that prostatic inflammation, perhaps triggered by MSU crystals, is a risk factor for prostate cancer.

We hypothesize that the deposition and accumulation of MSU crystals in the prostate (10) may induce a proinflammatory state in prostatic tissue with shared pathways to those in gout inflammation. Men with gout would have increased proclivity to respond to MSU crystals deposited in the prostate. *IL1R1* encodes an IL-1 receptor protein which binds IL-1α and IL-1β, and *IL1RN* encodes for an IL1 receptor antagonist (IL1-RA), which mediates IL-1α and IL-1β activity and inflammation. In eQTL analysis, rs9973741 (allele G, which is the risk allele for gout) associated with decreased expression of *IL1RN* in the prostate and decreased expression of both *IL1RN* and *IL1R1* in whole blood, while increasing *IL1RN* expression in the testes. This indicates that the putative pathway may involve complex patterns of regulation of gene expression. Further validation of our findings may lead to significant clinical treatments to mitigate the risk of prostate cancer, such as the use of urate lowering therapies to reduce prostatic inflammation. Notably allopurinol has previously been used to reduce symptoms of non-bacterial prostatitis (34, 35).

The exact mechanisms by which *SLC30A5* may contribute to a causal relationship is less clear. *SLC30A5* encodes for ZnT5 and transports zinc into membrane-bound organelles and out of the cell and is vital in maintaining zinc homeostasis (36). ZnT5 has been shown to be downregulated in prostate cancer tissue, and this downregulation is an early event in prostate cancer progression (33, 37).

Our study has various strengths. We utilized a two-sample MR analysis, which allowed for finding causal genetic relationships. We used SNPs representing specific aspects of gout risk and SU level, rather than using all SNPs which were significantly associated with gout, which allowed us to test specific mechanistic hypotheses. We included only men in the various genetic datasets used in our study. We tested the causal associations using three methods, IVW, weighted median, and MR-Egger, which robustly test for causal relationships and test for horizontal pleiotropy. However, our study also has limitations. We only included European men in our analysis, which means that these results may not be applicable to other populations. Even though we conducted tests for horizontal pleiotropy, the MR-Egger test does not guarantee that no pleiotropy exists, and we did not conduct extensive checks for vertical pleiotropy. Owing to lack of independent datasets, we were unable to include replication in the study design. Additionally, our study used genetic predictors of SU levels rather than of prostatic urate levels. The collective evidence supports that the prostate produces urate, but genetic predictors of serum urate levels may not reflect those of prostate urate level.

In conclusion, our MR analyses of a European male population provided evidence for a causal relationship between the non-hyperuricemia compartment of gout and prostate cancer, but not between SU and prostate cancer. These results suggest that clinical intervention to reduce gout-caused inflammation may reduce prostate cancer risk.

## Data Availability

All data produced are available online on the GWAS Catalog, available at https://www.ebi.ac.uk/gwas/.

## Grant Support/Acknowledgments

JAS is supported by research grants from the Patient-Centered Outcomes Research Institute (PCORI; CER-2020C1-19193; SDM-2017C2-8224) and the National Institutes of Arthritis, Musculoskeletal and Skin Diseases (NIAMS; P50 AR060772). JAS is also supported by the resources and the use of facilities at the VA Medical Center at Houston, Texas, and by the Center for Innovations in Quality, Effectiveness and Safety (#CIN 13-413), Houston, Texas.

