## Supplemental Figures for "Testing for Causal Association between Serum Urate, Gout, and Prostatic Cancer in European Males"

**Supplementary Figure 1:** Forest plot of all non-hyperuricemia compartment of gout SNPs to prostate cancer

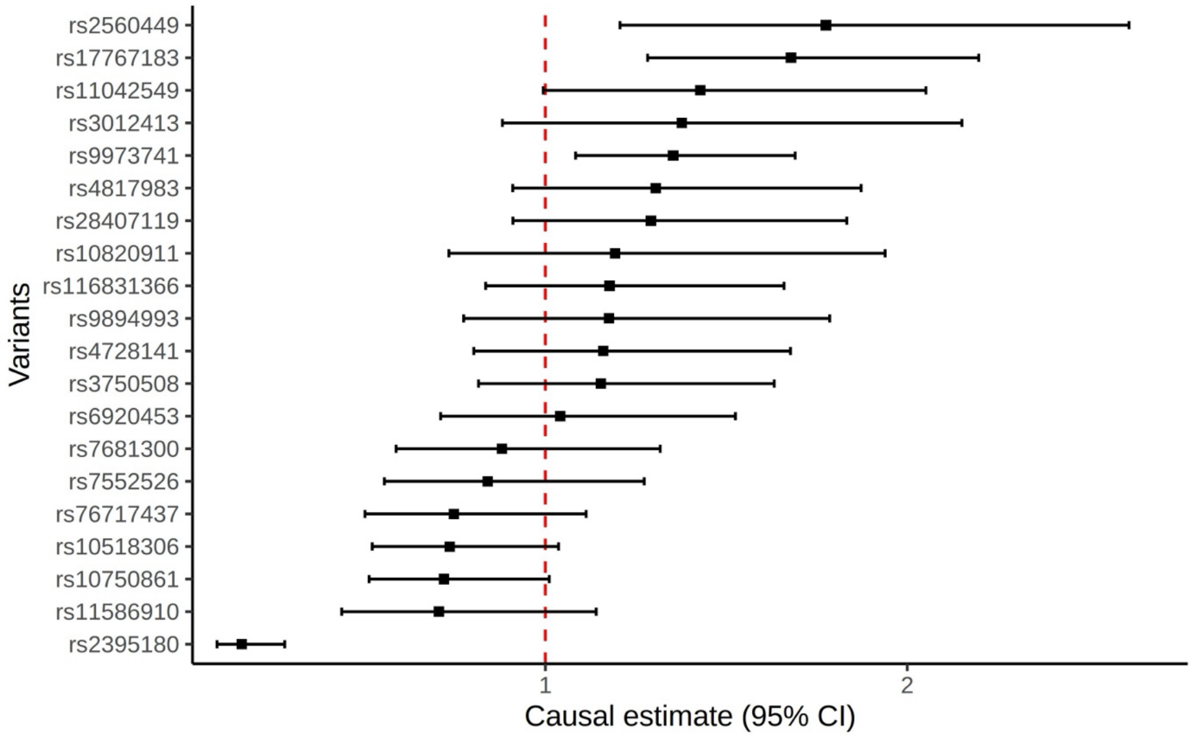

**Supplementary Figure 2:** LocusZoom plot of rs2560449 in prostate cancer (A) and in gout (B) in men.

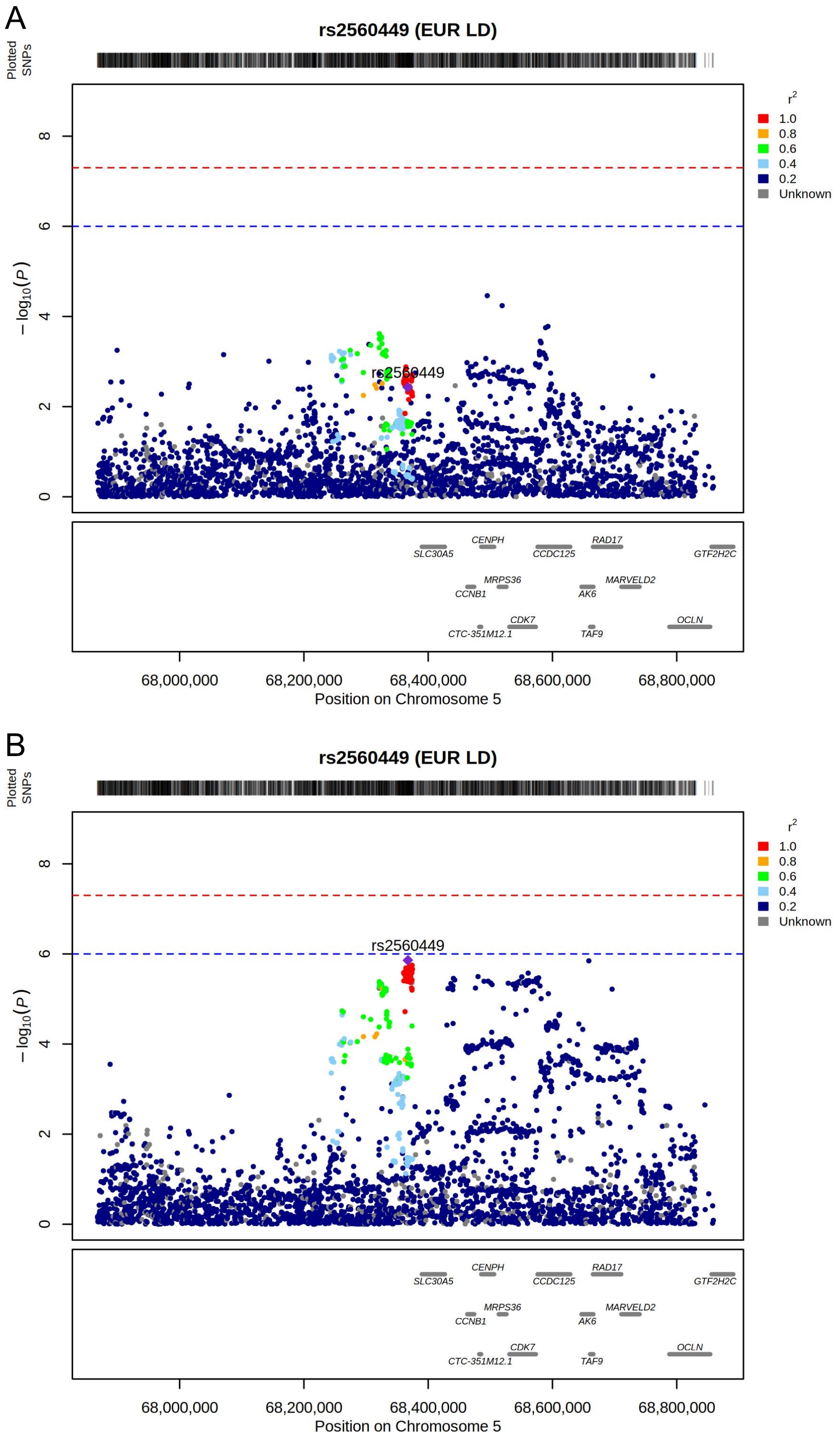

**Supplementary Figure 3:** LocusZoom plot of rs17767183 in prostate cancer (A) and in gout (B) in men.

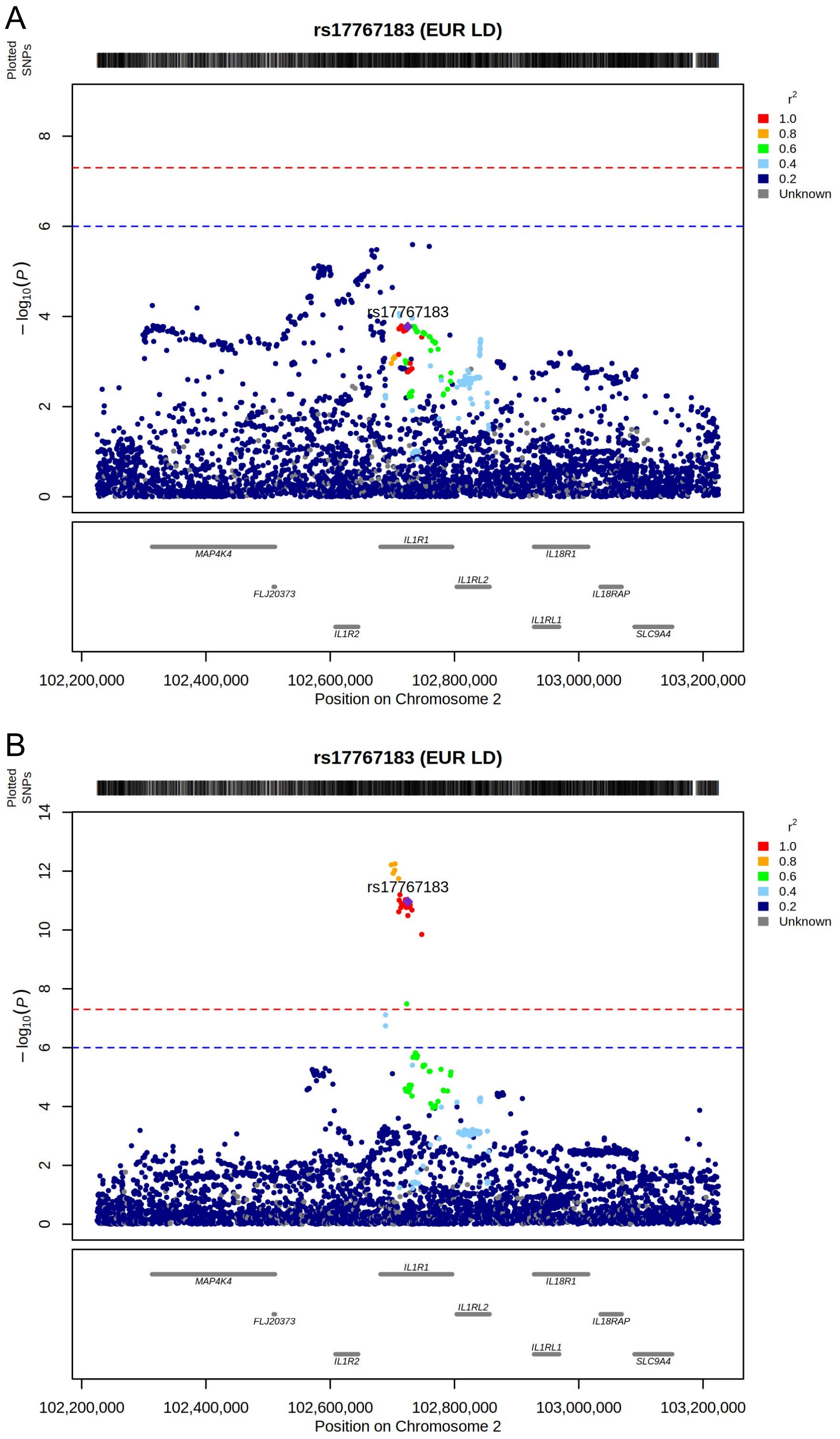

**Supplementary Figure 4:** LocusZoom plot of rs9973741 in prostate cancer (A) and in gout (B) in men.

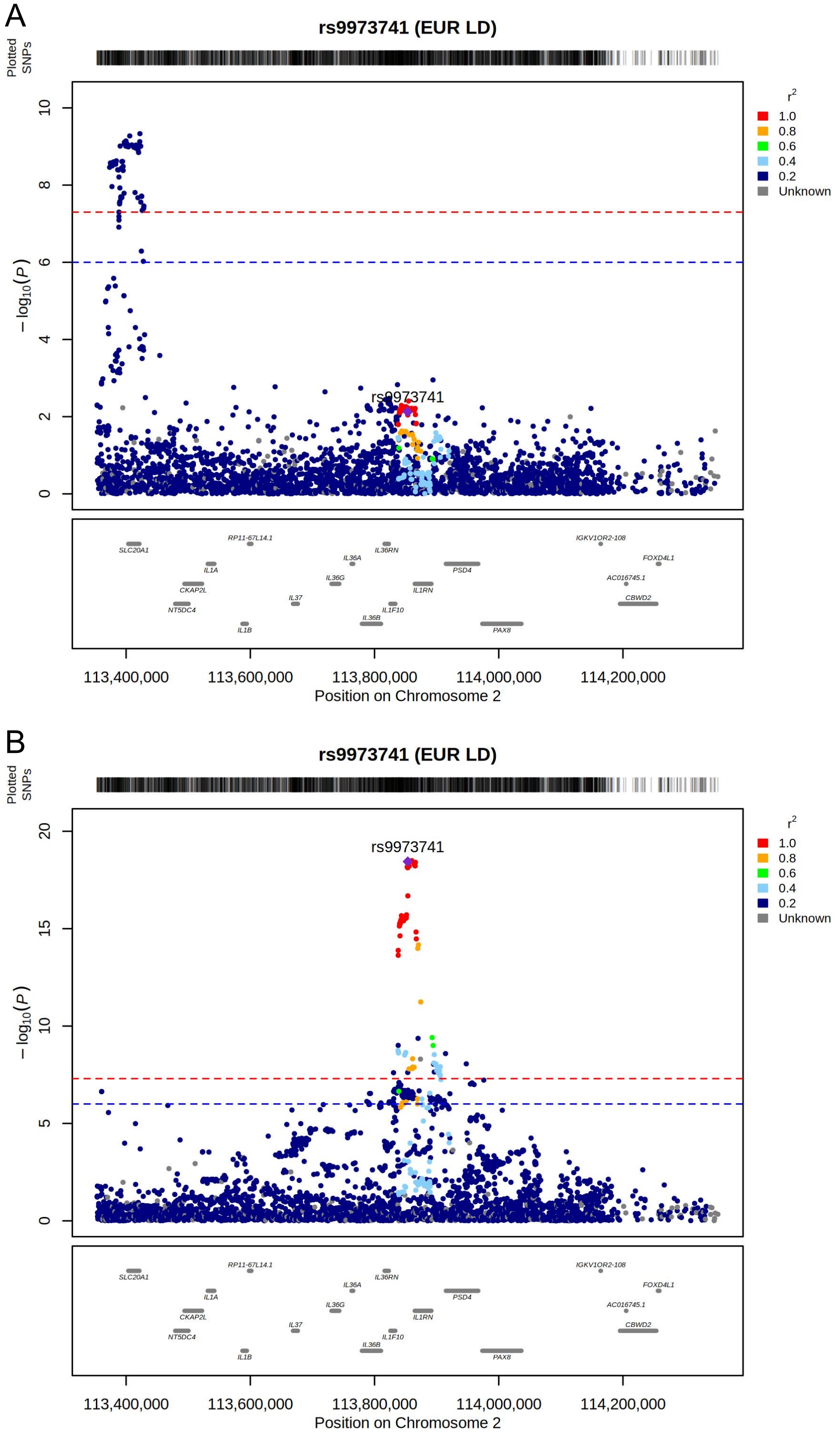

**Supplementary Figure 5:** LocusZoom plot of rs2395180 in prostate cancer (A) and in gout (B) in men.

A

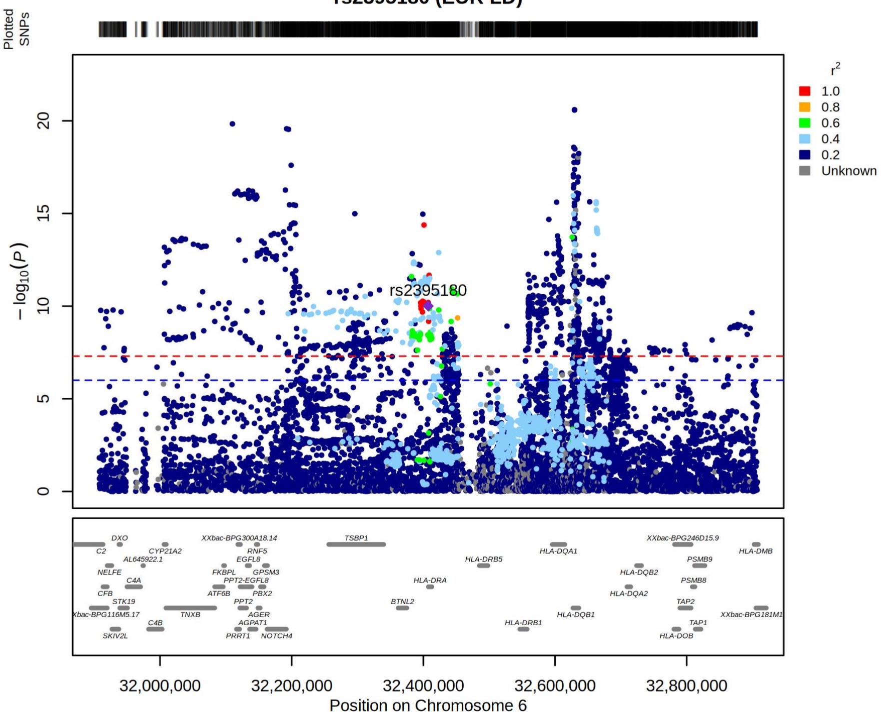


B

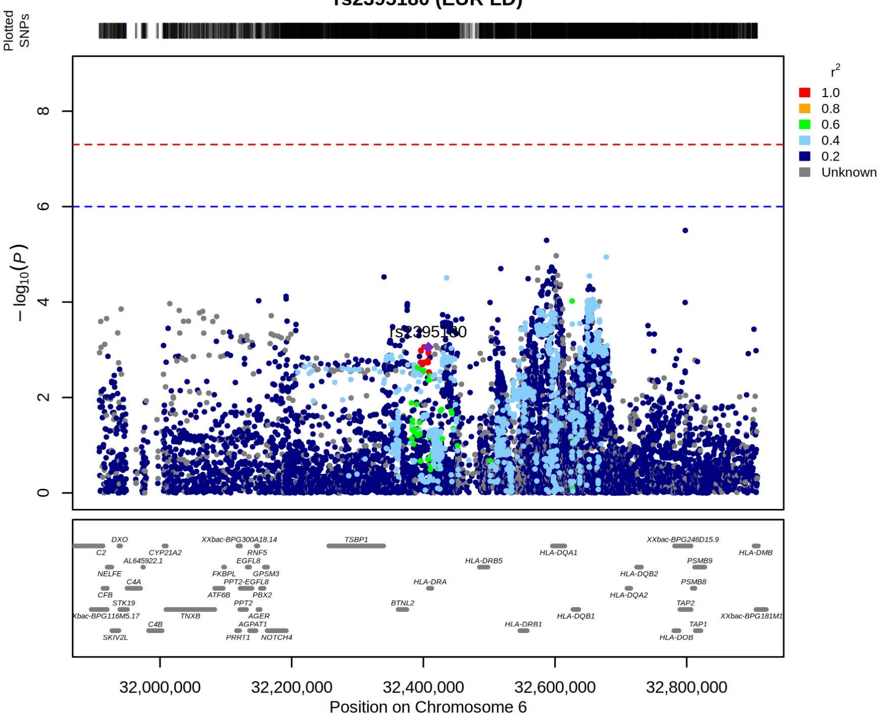

**Supplementary Figure 6:** LocusZoom of the eQTL for the association of rs9973741 and IL1RN in testis tissue (A) and with gout (B) and prostate cancer (C)

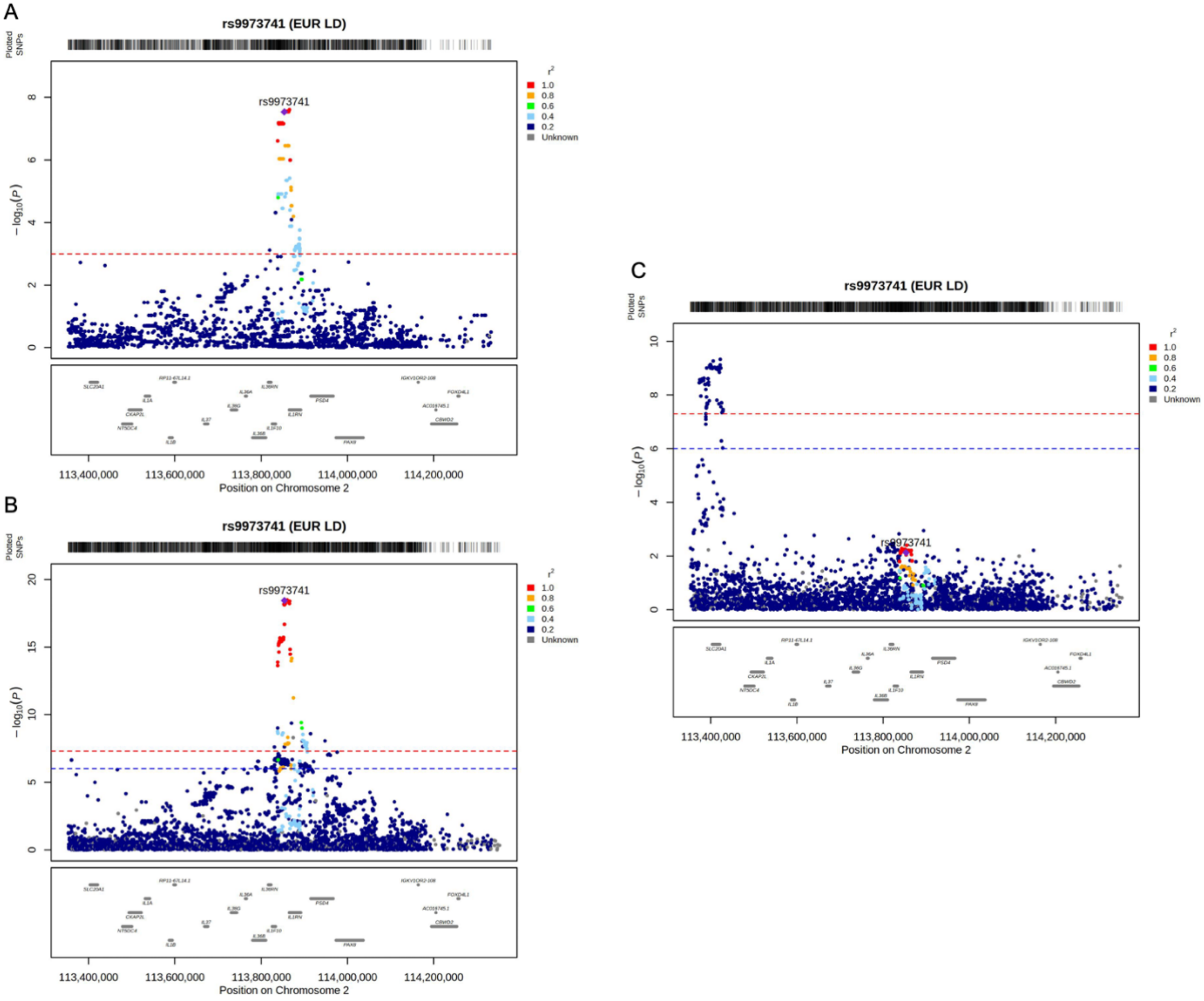

**Supplementary Figure 7:** Forest plot of all urate transporter SNPs to prostate cancer

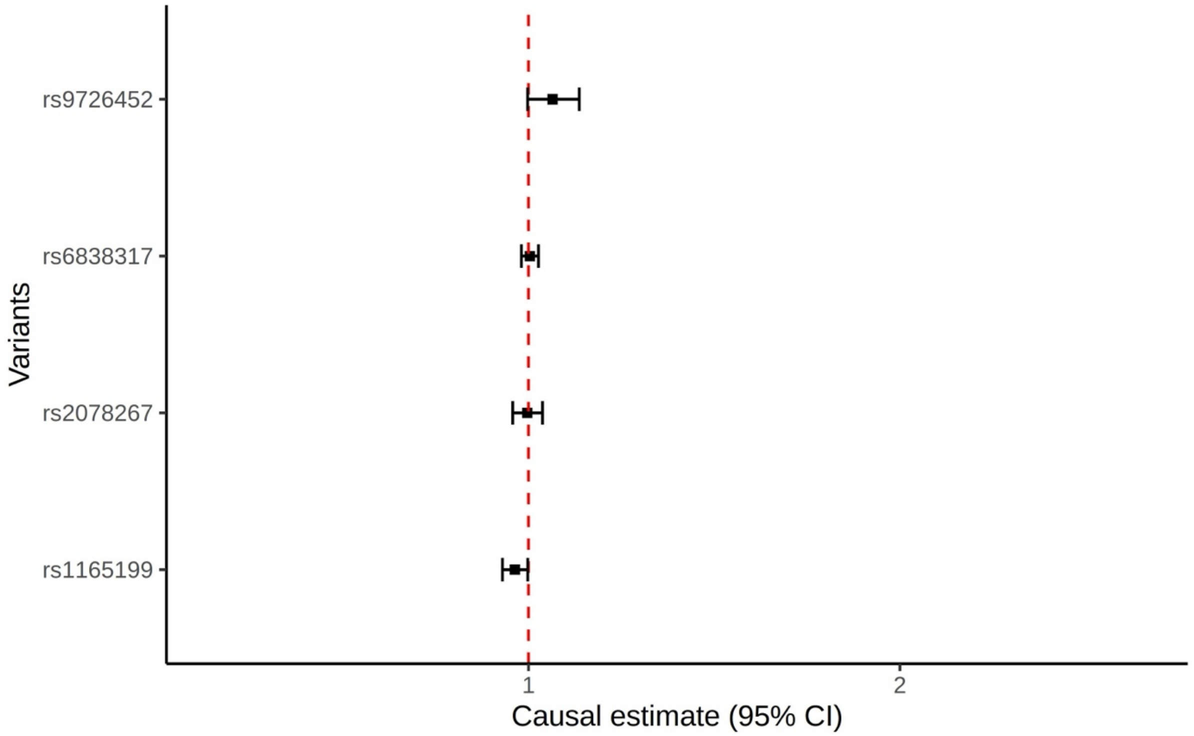
